# A Design Thinking Approach to Enhance Interprofessional Education Between Technology and Medicine for Innovative Elder Pain Care

**DOI:** 10.1101/2021.07.06.21260007

**Authors:** Patama Gomutbutra, Noppon Choosri, Peerasak Lerttrakarnnon

## Abstract

**Background:** Design thinking (DT) describes three stages in the design thinking cycle: 1) inspiration, which embodies the initial problem or opportunity; 2) ideation, which encompasses the development and refinement of ideas; and 3) implementation, which involves the introduction and application of the derived solution.

**Method:** The prospective educational program evaluation July 2020 – December 2020. A 150 min interactive workshop regarding developing innovative elder pain care. Medical students will be grouping into four medical students: 1 technological student. Then they will be assigned to match with a technology student. The interprofessional education design thinking activity consists of three parts; a brief introduction, the brainstorming process to identified pain points by persona and user journey to generate the idea, and the prototype presentation.

**Results:** Forty sixth-year medical students and twelve technological students participated. 58% of medical students and 95% of technological students perceived DT as very helpful for their careers. However, only 30% of medical students and 60% of technological students were inspired to develop actual prototypes after the course. Nearly all of the students responded they comfortable with this interdisciplinary project-based style. Still, the time of the workshop is too limited, and among those responding with no interest pursue the project gave a reason that they lack time after rotating to other courses.

**Conclusion:** Most medical students and technological students perceived DT as beneficial for their career and displayed satisfy in this co-project style. However, finding matched schedule of two disciplines made long-term interpersonal education challenging.

## Introduction

The rapid changing and complex society need a new generation who has 21st-century skill. The future physician is who both master the analytic thinking regarding clinical content and creative thinking as well as technology literacy. Interprofessional education (IPE) between technology and medicine has been growing in evidence in the past decades.

Design thinking (DT) is a framework for complex problem solving applied widely by various disciplines and recently emerging within healthcare. As a methodology, the origin of design thinking is often credited to Herbert A. Simon’s Sciences of the Artificial in 1969(1). The design process recently popularized by Tim Brown describes three stages in the design thinking cycle: 1) inspiration, which embodies the initial problem or opportunity; 2) ideation, which encompasses the development and refinement of ideas; and 3) implementation, which involves the introduction and application of the derived solution.

Previous medical, educational studies that applied DT showed that DT enhances understanding of complex issues and collaborative skills. A study integrated DT to medical ethic curriculum found students improved knowledge and attitudes toward organ transplantation (2). Another study applied DT to assign an interdisciplinary project for medicine and other science students to improve the health care environment. The results showed positive feedback in terms of increasing motivation and creativity capability (3). However, its use in Thailand’s medical education has never been addressing. We choose this elder pain topic because our team is also working on the project pain assessment by facial recognition technology (4)

## Methods

Study Population and Sampling:

1. The final year medical students rotating to the Faculty of Family Medicine during the four months study period (July 2020 – December 2020) 40 students
2. The 3rd year technology student who select to the human-computer subject 12 students The medical students will be grouping into 3-4 people. Then they will be assigned to match with a technology student.

The interventions

The total 150 minutes interactive workshop including three parts;

1. 30 minutes introduction about the principle of design thinking and elderly pain care.
2. 60 minutes. Each group would brainstorm to produce a personal persona and user journey in order to identify problem and prototype ideation based on human-centered
3. 30 minutes for prototype presentation. Each group present in pitching style for 10 minutes for each group in a seminar.

### Outcome measurement

The general aim of the study is to explore the feasibility of integrating design thinking to enhance interprofessional education between technology and medicine students.

The short term objective is to gather data from a checklist-questionnaire specifically:

Do the students perceive this activity as relevant or useful to their profession? : very useful, somewhat useful, and not useful

Do this activity inspired students to develop their prototype after the course: interest versus no interest

Is the programming assignment optimum to the students’ capacity? : too difficult, optimal, and too easy.

The longer-term objective is to further develop the joint curriculum between technology and medicine in the future—the themes and quotes from student responses to open-ended questions.

1. Which part of an activity that you like the most and why?
2. Which part of the activity that you think needs improvement, and in what way?

The activities depicted in Figure 1 and were recorded by verbal permission of participants, which could be reached at https://www.youtube.com/watch?v=PXuZYk-WcNk

## Results

Forty sixth-year medical students and twelve technological students participated. Among medical students, 58% perceived DT as very helpful for their career, 40 % perceived it as somewhat useful, 2% responded that not useful and more suitable to be an elective rather than a core course. Meanwhile, among technological students, nearly all (95%) think this activity is very useful.

30% of medical students, 80% of technological students were inspired to develop real prototypes after the course. Among those responding with no interest pursue the project gave the reason that they lack time after rotating to other courses. Nearly all (98%) of students responded they comfortable with this project-based education style. The details of these quantitative data are described in Table1.

**Table 1:** Demographic data of students participate in the DT workshop.

| Demographic data | Medical students (N=40) | Technological student(N=12) |
| --- | --- | --- |
| Which part of an activity that you like the most and why? | 23 (22-24) | 21 (19-24) |
| Which part of the activity that you think needs improvement, and in what way? | 23 (57.5%) | 6 (50%) |
| Expected career after graduate |  |  |
| 1. Working in the government sector | 22 (55 %) | 4 (33.3 %) |
| 2. Self-employed/enterpreuneur | 18 (45%) | 8 (66.7%) |
| Do the students perceive this activity as relevant or useful to their profession? |  |  |
| 1. Very useful | 23 (58%) | 10 (95%) |
| 2. Somewhat useful | 16 (40%) | 2 (5%) |
| 3. Not useful | 1 (2%) |  |
| Do this activity inspired students to develop their prototype after the course |  |  |
| 1. Interest to pursue the project | 12 (30%) | 7 (60%) |
| 2. No interest | 28 (70%) | 5 (40%) |
| Is the programming assignment optimum to the students' capacity? |  |  |
| 1. Too difficult | 1(2%) | 0 |
| 2. Optimal | 39 (98%) | 12 (100%) |
| 3. Too easy | 0 | 0 |

The qualitative data as themes and quotes from student responses to design thinking workshop showed many students like doing persona but the time of brainstorming is too limited and among whom responding no interest pursue the project gave the reason that they lack time after rotating to other courses. The thematic quotes were summarized in table 2

**Table 2:** Demographic data of students participate in the DT workshop.

| Questions | Medical students | Technological student |
| --- | --- | --- |
| Which part of an activity that you like the most and why? | -Storytelling persona formation that provided a new experience and different point of viewing - human-centered rather than epidemiologic centered as usual evidence-based medicine approach | -workshop part because this part let me face the real user that helps me to know the real problem and can solve the real point.<br>- the session of brainstorming for making a persona/journal map, because my teammates and I can show individual thoughts and exchange the experience for doing best. |
| Which part of the activity that you think needs improvement, and in what way? | - Workshop part in limited time.<br>- Introduction: may provide more information that relates to the workshop since it is a very new kind of for medical student<br>- Should be a selective course rather than a core course. | -this activity should be increased duration if I have more time; the opinion and thought maybe shown and exchanged for planning and improving a persona.<br>- The project could not pursue the completed project because of lack of time after rotating to the next course. |

## Discussion

The majority of medical students and technological students perceived DT as useful for their career and displayed satisfy in this co-project style. However, finding matched schedule of two disciplines made long-term interpersonal education become challenging.

Design thinking (DT) has gained attention as a method suitable to 21^st^-century education to redesign curricula and encourage students to solve problems using creativity and collaboration. Our participated students are categorized in generation Z (who was born between 1995 to 2012). They commonly are described as ‘digital naïve’ based on growing up along with the internet and social media. Generation Z tends to be naturally independent learners who look up to their own authority. (5). This character may explain the survey found that more than half of medical students and technological students prefer self-employed or being entrepreneurs over working under a more hierarchical job as government officers. Although relatively short attention span and less face-to-face communication skills compared to previous generations, they desire to be active problem-solvers for the sake of society. This may explain why DT could fulfill both their weakness by teamwork activity and enhance their strength in terms of social responsibility mindset.

There is some limitation of this study. First, the COVID19 situation in early to mid-2020 caused the number of students able to participate in the workshop less than expected. The second and most important barrier is the structure of the medical school curriculum which each course has many instructors get involve. This makes it difficult to arrange a regular matching teaching schedule with the technological school side, which be under one main instructor, as many students feedback, they could not pursue their prototype idea due to lack of continuous support. Third, there is no yet outcome measurement that reflects the real academic achievement. The solution that able to solve the targeted problem or to improve the quality of patient’s care as described in the literature (6). Since this outcome is impossible to get in a short time, the next study should plan for a longer follow-up.

In conclusion, our lesson learned from this DT-IPE project is well accepted by both medical and technological. The more flexible medical curriculum schedule will enhance long-term collaborative projects.

## Data Availability

The data could be available on request.

## Ethical Considerations

Ethics approval: The Faculty of Medicine Chiang Mai university IRB (EXEMPTION 7462/2020). All participants provided verbal consent to survey and video recording.

## Supplement material

The video clip demonstrates the design thinking workshop activities https://www.youtube.com/watch?v=PXuZYk-WcNk

